# Peritoneal Metastasis Prediction in Gastric Cancer: A Machine Learning Approach

**DOI:** 10.1101/2025.04.11.25325702

**Authors:** Shayan Jalali, Katayoon Dadkhah, Mirhamid Mirsaeid Ghazi

## Abstract

**Objective:** Evaluate the predictive efficacy of six machine learning (ML) algorithms in identifying peritoneal metastasis in gastric cancer (GC) patients.

**Methods:** Data from 809 GC patients (712 non-metastasis, 97 metastasis) were split into training and test sets (80:20). Six ML models—Decision Trees (DT), K-Nearest Neighbors (KNN), Support Vector Machines (SVM), Naive Bayes (NB), Random Forest (RF), and Logistic Regression (LR)—were assessed for feature importance and predictive performance.

**Results:** Lymph node positivity, lymph nodes cleaned, invasion depth, lymphatic invasion, and node dissection extent were key predictors. Among inflammatory markers, PLR was significant (p = 0.018), while NLR was not (p = 0.121). RF achieved the highest accuracy (97%), followed by SVM and LR.

**Conclusion:** ML enables robust prediction of peritoneal metastasis, with RF demonstrating the best performance. These findings highlight ML’s role in risk stratification, though multi-center validation is required for clinical application.

## Introduction

Gastrointestinal cancers, particularly gastric cancer (GC) and colorectal cancer (CRC), are among the most prevalent and lethal malignancies worldwide. They impose a significant burden on global healthcare systems, with CRC ranking as the third most commonly diagnosed cancer and the second leading cause of cancer-related mortality in both industrialized and developing nations [1]. Similarly, GC is widely recognized as an aggressive and heterogeneous malignancy, ranking fourth in incidence and second in cancer-related mortality globally [2]. Although its incidence has declined in certain regions, GC remains a major contributor to global cancer mortality [1, 3]. Prognosis for patients diagnosed at advanced stages remains poor, primarily due to metastatic dissemination. Patterns of metastatic spread vary depending on the primary tumor origin [4]. In CRC, the liver is the predominant site of metastasis, occurring in approximately 50% of cases [5]. Similarly, GC frequently metastasizes to the liver and other distant organs, complicating clinical management and limiting therapeutic options [1]. However, the peritoneal membrane represents one of the most common metastatic sites in GC, significantly impacting patient prognosis [6]. Studies indicate that a substantial proportion of newly diagnosed GC cases present with malignant ascites, highlighting the severity and high prevalence of peritoneal metastasis—particularly in advanced stages [6]. Patients with peritoneal metastasis face markedly poor prognoses, with median survival ranging from four to six months. Furthermore, malignant ascites often leads to severe complications, including intestinal obstruction and renal insufficiency, significantly impairing patients’ quality of life [7, 8]. With advances in precision oncology, the need for accurate and reliable prognostic tools has become increasingly apparent [9]. In this context, machine learning (ML) has emerged as an asset in medical decision-making, enhancing predictive accuracy across multiple clinical domains [10, 11]. ML algorithms have demonstrated success in cancer prognosis and metastasis prediction, providing critical insights into disease progression and patient outcomes [12, 13]. Beyond oncology, ML has shown substantial efficacy in infectious disease modeling by effectively analyzing large-scale datasets, facilitating early detection, and optimizing treatment strategies [14, 15]. Despite advances in cancer diagnostics, accurate diagnosis of peritoneal metastasis—especially at early stages—remains a significant clinical challenge. Laparoscopy and pathological assessments are the gold-standard diagnostic methods; however, their invasive nature, associated procedural risks, and high costs limit their routine use in clinical practice [16, 17]. To address these constraints, recent research has integrated ML with medical imaging and clinical data, resulting in enhanced predictive accuracy for cancer diagnoses [18, 19]. For instance, radiomics-based texture analysis of CT images has demonstrated considerable promise in predicting occult peritoneal metastasis in advanced GC, offering a valuable non-invasive approach for early detection [19, 20]. Additionally, ML-driven predictive models have been developed to analyze metastatic patterns, facilitating personalized patient management and enhancing clinical decision-making [21, 22]. However, despite these promising developments, the application of ML specifically for predicting peritoneal metastasis in GC remains relatively underexplored. Although ML has proven successful in various oncology settings, its predictive efficacy in forecasting peritoneal recurrence warrants further exploration [23]. To bridge this gap, this study evaluates the predictive efficacy of six ML algorithms in identifying peritoneal metastasis among gastric cancer patients who underwent surgical treatment, with the goal of enhancing diagnostic precision and informing therapeutic strategies. By employing extensive datasets and advanced computational methods, this research aims to improve clinical management strategies and ultimately optimize outcomes for patients suffering from this highly challenging malignancy.

## Materials and Methods

### Participants

In this retrospective study, we initially reviewed clinical records of 1,199 patients diagnosed with gastric cancer (GC) who underwent surgical treatment. Before surgery, all participants underwent preoperative computed tomography (CT) scans to rule out peritoneal metastasis. After applying exclusion criteria, including previous gastrectomy, existing liver diseases (e.g., cirrhosis), history of other malignancies, severe bleeding or autoimmune diseases, receipt of preoperative chemoradiotherapy, severe inflammatory or hematological disorders, distant metastases outside the abdomen, and cases with missing or incomplete data, the final dataset comprised 809 patients. The demographic characteristics of the final cohort (n = 809) were as follows: The mean age was 63.7 years, with no significant difference between patients with (64.03 ± 11.61 years) and without (63.66 ± 11.21 years) peritoneal metastasis. Gender distribution included 550 males (77.2%) and 76 males (78.4%) in the non-metastasis and metastasis groups, respectively, and 162 females (22.8%) and 21 females (21.6%) in the respective groups, maintaining a similar gender ratio across groups. Further collected demographic and clinical data included body mass index (BMI) and American Society of Anesthesiologists (ASA) scores, indicating patients’ preoperative physical status. Additional data encompassed family medical history, tumor characteristics (location, size, histopathological differentiation, lymphatic invasion, and pathological type classified as ulcerative or non-ulcerative), and routine hematological indices, including neutrophils, lymphocytes, platelets, monocytes, Neutrophil-to-Lymphocyte Ratio (NLR), and Platelet-to-Lymphocyte Ratio (PLR). All diagnoses and pathological classifications were confirmed through histological examination.

### Study Measures

Study Measures encompass a range of demographic, clinical, and laboratory variables. Demographically, age was treated as a continuous variable with no significant difference noted between patients with and without peritoneal metastasis. Gender was categorized into male and female, with the study comprising 550 males (77.2%) and 162 females (22.8%) in the non-metastasis group and 76 males (78.4%) and 21 females (21.6%) in the metastasis group, reflecting a similar distribution between groups (P = 0.909). Body Mass Index (BMI) was also analyzed as a continuous variable. Clinically, the ASA Score was used to assess physical status in three levels, indicating preoperative health, with most patients classified under level II (85.3% in the non-metastasis group vs. 85.6% in the metastasis group, P = 0.969) for moderate systemic disease. TNM Stage classified the extent of cancer spread from I to IV, aiding in understanding disease progression (TNM Stage III: 51.1% in the non-metastasis group vs. 50.5% in the metastasis group, P = 0.833). Histology Type was determined based on cellular composition and appearance, influencing treatment response (Type 1: 10.1% in the non-metastasis group vs. 3.1% in the metastasis group, Type 2: 18.3% vs. 9.3%, Type 3: 51.0% vs. 62.9%, Type 4: 20.6% vs. 24.7%, P = 0.009), while Borrmann Type was classified based on the gross anatomical appearance of gastric cancer, indicating tumor aggressiveness (Type 1: 4.5% in the non-metastasis group vs. 7.2% in the metastasis group, Type 2: 88.9% vs. 79.4%, Type 3: 6.6% vs. 13.4%, P = 0.023). Pathology Type was divided into ulcerative and non-ulcerative, describing tumor growth characteristics and prognostic implications (non-ulcerative: 11.2% in the non-metastasis group vs. 19.6% in the metastasis group; Ulcerative: 88.8% vs. 80.4%, P = 0.029). Invasion Depth was categorized into three levels to reflect the penetration depth of tumor cells (T3/T4: 63.9% in the non-metastasis group vs. 94.8% in the metastasis group, P < 0.001), and Grouped Invasion Depth simplified these categories for analysis. Lymph Nodes were assessed through Lymph Nodes Cleaned and Lymph Nodes Positive measures, offering insights into the extent of cancer spread and surgical thoroughness (Lymph Nodes Cleaned: 19.53 ± 10.12 in the non-metastasis group vs. 16.76 ± 9.74 in the metastasis group, P = 0.011; Lymph Nodes Positive: 4.21 ± 6.11 in the non-metastasis group vs. 8.29 ± 7.03 in the metastasis group, P < 0.001). Lymphatic Invasion indicated metastatic potential into the lymphatic system. Laboratory measures included PLR and NLR as markers of systemic inflammation associated with cancer prognosis, Hemoglobin PreOp as an indicator of general health and surgical risk, and various blood counts like Platelet Count, Albumin Level, Neutrophil Count, Lymphocyte Count, Monocyte Count, and WBC Count, which provide a comprehensive picture of the patient’s immune status and inflammatory response, crucial in cancer management.

### Statistical Analysis

All statistical analyses were conducted using R software (version 4.4). Continuous variables were expressed as mean ± standard deviation (SD) and compared using the t-test, while categorical variables were presented as counts (n) and percentages (%), analyzed with the χ^2^ test. A p-value < 0.05 was considered statistically significant. To account for missing data, multiple imputation techniques were applied, ensuring data integrity and minimizing potential biases. The dataset was randomly stratified into training and test sets using an 80:20 split to train and validate predictive models [24]. Data normalization and the construction of prognostic weights were performed before model training [25]. Correlation analysis was conducted to identify relationships between variables and determine key predictive factors influencing peritoneal metastasis [26]. To develop robust predictive models, six machine learning (ML) algorithms were implemented: Decision Trees (DT), K-Nearest Neighbors (KNN), Support Vector Machines (SVM), Naive Bayes (NB), Random Forest (RF), and Logistic Regression (LR). Each model was evaluated based on its ability to handle large datasets efficiently and enhance predictive accuracy by identifying the most influential variables [27]. Model performance was rigorously assessed using multiple evaluation metrics, including Accuracy, Sensitivity (True Positive Rate), Specificity (True Negative Rate), Positive Predictive Value (PPV), Negative Predictive Value (NPV), Mean Squared Error (MSE), and Area Under the Curve (AUC). Accuracy was defined as the ratio of correctly classified samples to the total number of cases. Sensitivity and Specificity measured the model’s ability to correctly identify positive and negative outcomes, respectively, while PPV and NPV indicated the probability that patients with positive and negative test results had or did not have peritoneal metastasis [28]. MSE was utilized to assess the average squared differences between observed actual outcomes and model predictions, where lower values indicated superior model performance. To enhance model generalizability and prevent overfitting, a 5-fold cross-validation strategy was applied [29]. Additionally, regularization techniques and hyperparameter tuning were implemented to optimize model parameters, improving predictive accuracy while maintaining model stability [30]. This comprehensive analytical approach ensures that the developed machine learning models are both robust and reliable in predicting peritoneal metastasis in gastric cancer patients.

### Data Availability

The dataset supporting the findings of this study is publicly available in the Biostudies database under accession number S-EPMC5383064. It can be accessed at https://www.ebi.ac.uk/biostudies/studies?query=S-EPMC5383064. The dataset comprises comprehensive clinical records of 1,199 gastric cancer (GC) patients who underwent surgical treatment.

## Results

Our retrospective study analyzed 809 patients treated for gastric cancer, with a male predominance of 77.4%, including 626 men and 183 women. No significant age differences were observed between patients with and without peritoneal metastases, averaging 64.03 ± 11.61 years and 63.66 ± 11.21 years, respectively (p = 0.76). A statistically significant difference in Body Mass Index (BMI) was noted; patients who developed peritoneal metastases had a slightly lower BMI (21.09 ± 3.18 vs. 21.82 ± 3.09 kg/m^2^, p = 0.03). Key indicators for peritoneal metastasis included a significantly higher Platelet-to-Lymphocyte Ratio (PLR) in patients with metastases compared to those without (168.15 ± 76.30 vs. 149.08 ± 74.37, p = 0.018), whereas the Neutrophil-to-Lymphocyte Ratio (NLR) did not show a significant difference (2.85 ± 1.64 vs. 2.58 ± 1.58, p = 0.121). These markers reflect the inflammatory state and tumor microenvironment, influencing the likelihood of metastatic spread. Further, Hemoglobin PreOp levels were notably lower in patients with peritoneal metastases (112.51 ± 22.88 vs. 117.95 ± 23.98, p = 0.035), suggesting anemia associated with more aggressive disease states. Critical clinical features strongly associated with peritoneal metastasis included the extent of lymph nodes cleaned (16.76 ± 9.74 vs. 19.53 ± 10.12, p = 0.011), number of lymph nodes positive (8.29 ± 7.03 vs. 4.21 ± 6.11, p < 0.001), presence of lymphatic invasion (p < 0.001), and extent of node dissection (p < 0.001). Additionally, invasion depth was a significant predictor of metastasis, with 94.8% of metastatic cases classified as T3/T4 compared to 63.9% in non-metastatic cases (p < 0.001). These findings underscore the importance of these variables in predicting peritoneal metastasis, helping clinicians identify patients at higher risk and tailor their management strategies more effectively.

**Table 1.**
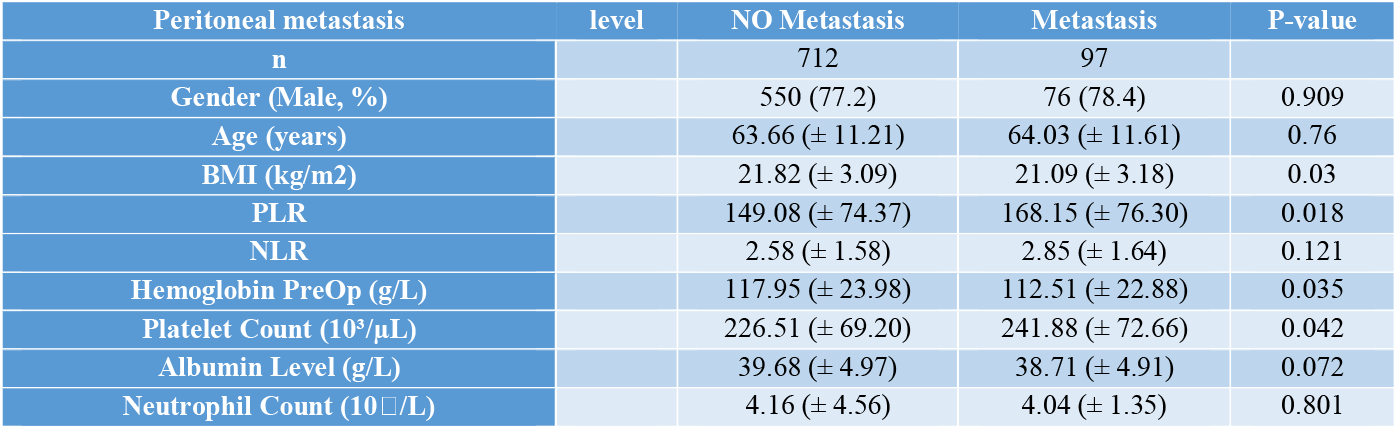
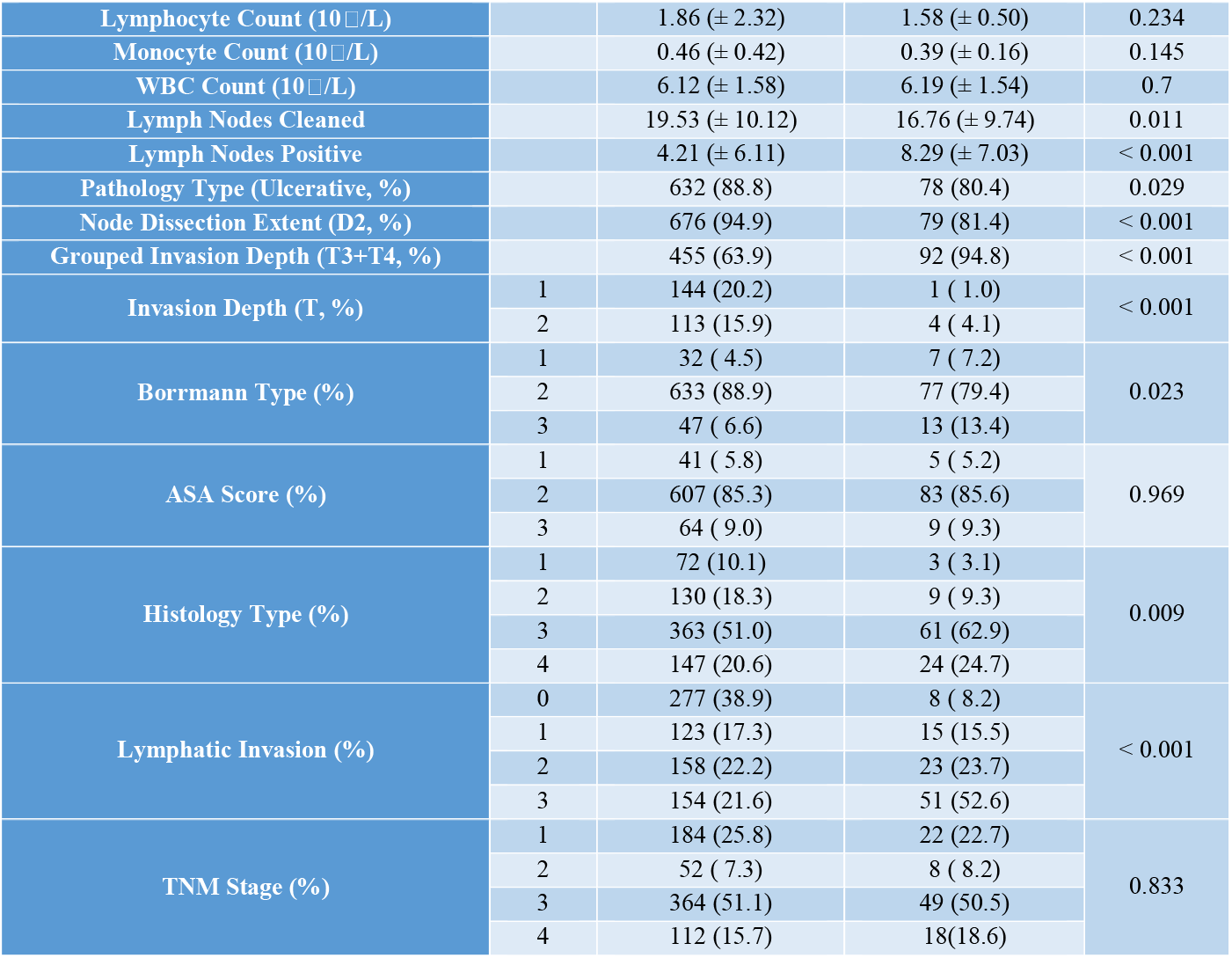
Baseline Data. *NLR, neutrophil-tolymphocyte ratio; PLR, platelet-to-lymphocyte ratio; WBC, white blood cell; Hemoglobin PreOp, hemoglobin levels in a patient’s blood prior to undergoing surgery*

### Classification Performance

The predictive performance of six machine learning (ML) models—K-Nearest Neighbors (KNN), Support Vector Machines (SVM), Naive Bayes (NB), Decision Trees (DT), Random Forest (RF), and Logistic Regression (LR)—was evaluated based on key performance metrics, including sensitivity, specificity, accuracy, Positive Predictive Value (PPV), and Negative Predictive Value (NPV). Among these models, Random Forest (RF) emerged as the top performer, demonstrating the highest accuracy, superior PPV, and excellent NPV, making it particularly reliable for both identifying and excluding peritoneal metastasis. Logistic Regression (LR) exhibited balanced performance across all metrics, though with a slightly lower PPV, positioning it as a strong and stable predictive model. Support Vector Machines (SVM) showed strong specificity, making it particularly effective in predicting true positives with minimal false positives. Naive Bayes (NB) stood out for its high sensitivity and the highest NPV, excelling in correctly identifying negative cases while minimizing false negatives. K-Nearest Neighbors (KNN) demonstrated high specificity, though its lower PPV indicated a higher likelihood of false positives. Decision Trees (DT), while having lower sensitivity, maintained high specificity and a reasonable PPV, contributing to its role as a useful classifier despite its comparatively weaker sensitivity. This comparative analysis highlights that Random Forest, Logistic Regression, and Naive Bayes provide robust predictive performance, each excelling in different aspects of accuracy and reliability. The optimal choice among these models depends on specific clinical objectives, such as minimizing false negatives to ensure early detection or reducing false positives to avoid unnecessary interventions.

**Figure 1.**
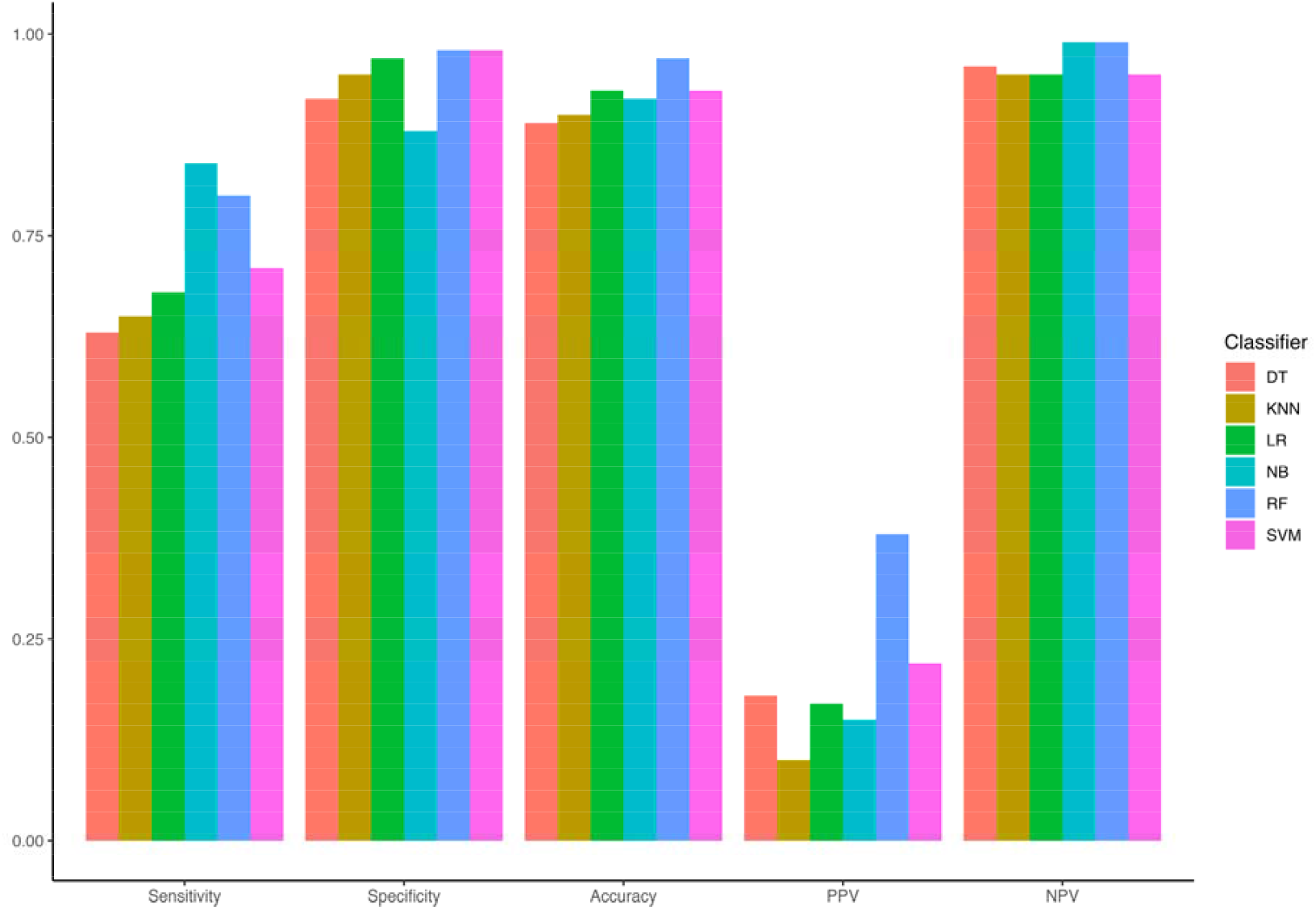
Comparison of Classifier Performance - This figure illustrates the performance metrics such as Sensitivity, Specificity, Accuracy, PPV, and NPV for each classifier used in the study.

### Feature Importance Analysis

To identify the most influential predictors of peritoneal metastasis in gastric cancer patients, Percentage Increase in Mean Squared Error (%IncMSE) was used as the primary metric. The analysis revealed that ‘Lymph Nodes Positive’ exhibited the highest %IncMSE, indicating its strong impact on model accuracy. Other key predictors included ‘Lymph Nodes Cleaned’, ‘Lymphatic Invasion’, ‘Node Dissection Extent’, and ‘Grouped Invasion Depth’, all of which significantly influenced classification performance. Additionally, ‘Invasion Depth’, ‘Neutrophil-to-Lymphocyte Ratio (NLR)’, ‘Platelet-to-Lymphocyte Ratio (PLR)’, ‘Albumin Level’, and ‘Borrmann Type’ were identified as important variables, reinforcing their relevance in predicting peritoneal metastasis. Beyond clinical and pathological factors, demographic features such as Age and BMI were also assessed. While they exhibited some level of significance, their primary value lies in providing clinical context, contributing to a broader understanding of disease progression and patient risk stratification. These findings underscore the importance of integrating clinical, pathological, and demographic factors to enhance the predictive power of machine learning models in assessing peritoneal metastasis risk.

**Figure 2.**
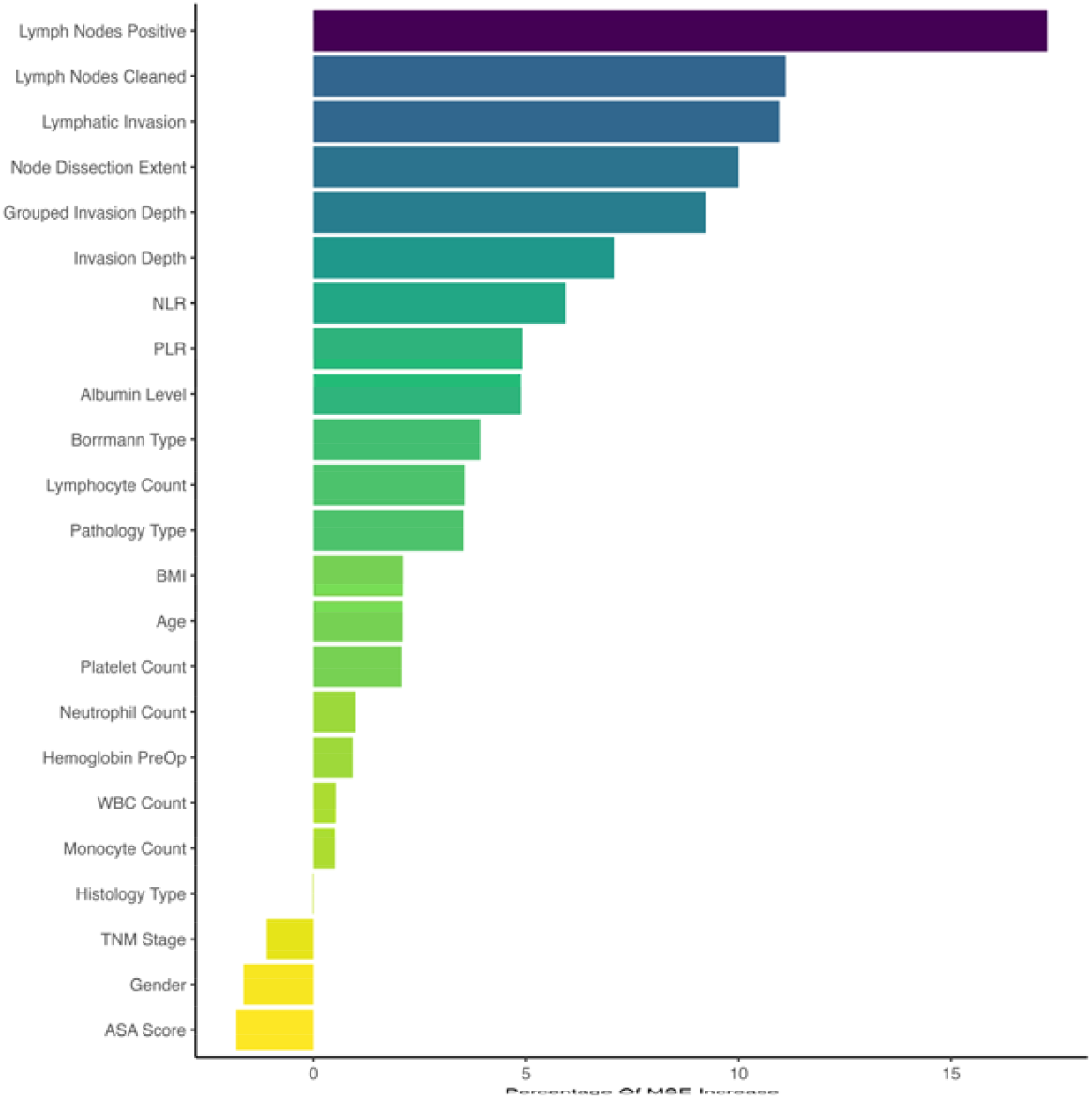
Variable Importance of Features Included in Machine Learning Algorithm for Prediction of Peritoneal Metastasis

## Discussion

Our study explores the complexities of predicting peritoneal metastasis in gastric cancer patients using machine learning techniques, highlighting the significance of several predictive variables. Gastric cancer remains a leading cause of cancer-related morbidity and mortality worldwide [2]. Peritoneal metastasis represents a major challenge in postoperative prognosis due to its location within the peritoneum—a mesothelial cell-lined membrane that encapsulates the abdominopelvic cavity. This anatomical feature frequently becomes a site for malignant tumor dissemination, significantly affecting patient survival and quality of life [31-33]. In our analysis, machine learning models demonstrated high predictive accuracy for peritoneal metastasis, with the Random Forest (RF) model achieving an accuracy of 97%, making it the most effective classifier. Other models, including Support Vector Machines (SVM) and Logistic Regression (LR), also exhibited strong performance. These findings reinforce the clinical potential of machine learning, providing a non-invasive, data-driven approach for the early detection of peritoneal metastasis [34]. Feature selection plays a crucial role in enhancing ML-based predictions, as demonstrated in various studies related to metastatic cancer progression [35]. The integration of clinical and laboratory features into ML models has been widely validated, particularly in fields ranging from stroke prediction to cancer metastasis assessment, underscoring the necessity of a comprehensive feature selection approach [33]. In our study, key predictive variables included lymph node positivity, invasion depth, and inflammatory markers, which significantly contributed to model performance. Selecting the most relevant features not only improves accuracy but also enhances model interpretability, making it more clinically applicable. Particularly, factors such as lymph node involvement, invasion depth, and inflammatory markers like the Platelet-to-Lymphocyte Ratio (PLR) and Neutrophil-to-Lymphocyte Ratio (NLR) were identified as critical variables influencing the prediction outcomes. These elements reflect the aggressive nature of the disease and its systemic impact, underscoring the need for effective predictive models in clinical settings. However, while PLR was significantly higher in patients with peritoneal metastasis (p = 0.018), NLR did not show a significant difference (p = 0.121), suggesting that PLR may be a stronger inflammatory predictor in this context. Our findings align with previous research indicating that increased PLR and NLR are associated with worse prognostic outcomes in gastric cancer and other malignancies [36, 37]. Additionally, our study highlights the prognostic importance of TNM staging, particularly in advanced-stage patients (III and IV), where peritoneal metastasis risk is significantly higher. Specific pathological factors, such as Borrmann Type 2 and 3, also emerged as strong predictors of peritoneal dissemination, aligning with existing literature that associates these subtypes with poor survival outcomes [38, 39]. In our dataset, peritoneal metastasis was detected in 22 (22.7%) of Stage I patients, 8 (8.2%) of Stage II patients, 49 (50.5%) of Stage III patients, and 18 (18.6%) of Stage IV patients, reflecting an increasing trend of metastasis with cancer progression, despite the overall p-value (p = 0.833) not reaching statistical significance. Similarly, Borrmann Type classification indicated that peritoneal metastasis was present in 7 (7.2%) of Type 1 cases, 77 (79.4%) of Type 2 cases, and 13 (13.4%) of Type 3 cases (p = 0.023). These findings suggest a potential clinical trend that warrants further investigation in larger, multi-institutional cohorts. In addition to oncological factors, nutritional status, particularly BMI, was also associated with peritoneal metastasis risk. Previous studies have reported that low BMI correlates with poorer overall survival and increased postoperative complications in gastric cancer [40, 41]. Our findings align with this, as patients with peritoneal metastasis had a significantly lower BMI than those without (p = 0.03), reinforcing the role of nutritional status as a potential prognostic indicator. Despite the promising results, our study has some limitations. The retrospective nature of the analysis and reliance on postoperative data restricts the applicability of the findings to preoperative settings, where predictive accuracy is crucial for planning treatment strategies. Moreover, the study’s single-institution scope may limit the generalizability of the results, necessitating validation through prospective, multicenter studies to confirm the robustness and applicability of the predictive models developed. Additionally, previous research has demonstrated that ML models can integrate heterogeneous data sources, such as physiological signals, biomarkers, and imaging data, to enhance predictive accuracy in clinical applications. These findings align with our study, emphasizing the importance of incorporating diverse clinical-pathological factors to improve prediction models [35]. In conclusion, while our study advances the understanding of factors influencing peritoneal metastasis in gastric cancer, it also highlights the potential of machine learning in enhancing prognostic assessments. Future research should aim to refine these models with a broader set of clinical-pathological factors and validate their predictive power in larger, diverse patient cohorts.

## Conclusion

This study demonstrated that Random Forest (RF) achieved the highest accuracy (97%) in predicting peritoneal metastasis in gastric cancer, followed by SVM and LR. Lymph node positivity, lymph nodes cleaned, invasion depth, lymphatic invasion, and node dissection extent were the most influential predictive factors. Among inflammatory markers, PLR was significant, while NLR was not. Advanced TNM stages (III & IV) and Borrmann Type 2 & 3 tumors were strongly associated with metastasis. Additionally, lower BMI and hemoglobin levels were observed in metastatic cases. Despite high model accuracy, multi-center validation is needed to confirm clinical applicability.

## References

1. Dong, D., et al., Deep learning radiomic nomogram can predict the number of lymph node metastasis in locally advanced gastric cancer: an international multicenter study. Annals of Oncology, 2020. 31(7): p. 912–920.

2. Ong, H.S. and B.J.C.R.A.-P. Mark Smithers, Epidemiology of gastric cancer. 2004. 2(01): p. 1–7.

3. An, C., et al., Deep learning radiomics of dual-energy computed tomography for predicting lymph node metastases of pancreatic ductal adenocarcinoma. 2022: p. 1–13.

4. Soltanyzadeh, M., et al., Clarifying differences in gene expression profile of umbilical cord vein and bone marrow-derived mesenchymal stem cells; a comparative in silico study. Informatics in Medicine Unlocked, 2022. 33: p. 101072.

5. Li, Y., et al., Establishment of a new non-invasive imaging prediction model for liver metastasis in colon cancer. 2019. 9(11): p. 2482.

6. Maeda, H., M. Kobayashi, and J.J.W.J.o.G.W. Sakamoto, Evaluation and treatment of malignant ascites secondary to gastric cancer. 2015. 21(39): p. 10936.

7. Haghzad, T., et al., A computational approach to assessing the prognostic implications of BRAF and RAS mutations in patients with papillary thyroid carcinoma. Endocrine, 2024. 86(2): p. 707–722.

8. Houri, H., et al., High prevalence rate of microbial contamination in patient-ready gastrointestinal endoscopes in Tehran, Iran: an alarming sign for the occurrence of severe outbreaks. Microbiology Spectrum, 2022. 10(5): p. e01897–22.

9. Zareei, S., et al., PeptiHub: a curated repository of precisely annotated cancer-related peptides with advanced utilities for peptide exploration and discovery. Database, 2024. 2024: p. baae092.

10. Irankhah, L., et al., Analyzing the performance of short-read classification tools on metagenomic samples toward proper diagnosis of diseases. Journal of bioinformatics and computational biology, 2024. 22(5): p. 2450012.

11. Kharaghani, A.A., et al., High prevalence of Mucosa-Associated extended-spectrum β-Lactamase-producing Escherichia coli and Klebsiella pneumoniae among Iranain patients with inflammatory bowel disease (IBD). Annals of Clinical Microbiology and Antimicrobials, 2023. 22(1): p. 86.

12. Khorsand, B., et al., OligoCOOL: a mobile application for nucleotide sequence analysis. Biochemistry and Molecular Biology Education, 2019. 47(2): p. 201–206.

13. Khorsand, B., et al., Alpha influenza virus infiltration prediction using virus-human protein-protein interaction network. Mathematical Biosciences and Engineering, 2020. 17(4): p. 3109–3129.

14. Khorsand, B., A. Savadi, and M. Naghibzadeh, Comprehensive host-pathogen protein-protein interaction network analysis. BMC bioinformatics, 2020. 21: p. 1–22.

15. Khorsand, B., A. Savadi, and M. Naghibzadeh, Parallelizing assignment problem with DNA strands. Iranian Journal of Biotechnology, 2020. 18(1): p. e2547.

16. Khorsand, B., A. Savadi, and M. Naghibzadeh, SARS-CoV-2-human protein-protein interaction network. Informatics in medicine unlocked, 2020. 20: p. 100413.

17. Khorsand, B., et al., Overrepresentation of Enterobacteriaceae and Escherichia coli is the major gut microbiome signature in Crohn’s disease and ulcerative colitis; a comprehensive metagenomic analysis of IBDMDB datasets. Frontiers in cellular and infection microbiology, 2022. 12: p. 1015890.

18. Dong, D., et al., Development and validation of an individualized nomogram to identify occult peritoneal metastasis in patients with advanced gastric cancer. 2019. 30(3): p. 431–438.

19. Liu, S., et al., Radiomics analysis using contrast-enhanced CT for preoperative prediction of occult peritoneal metastasis in advanced gastric cancer. 2020. 30: p. 239–246.

20. Kim, H.Y., et al., Could texture features from preoperative CT image be used for predicting occult peritoneal carcinomatosis in patients with advanced gastric cancer? 2018. 13(3): p. e0194755.

21. Shinagare, A.B., et al., High-grade serous ovarian cancer: use of machine learning to predict abdominopelvic recurrence on CT on the basis of serial cancer antigen 125 levels. 2018. 15(8): p. 1133–1138.

22. Tahmassebi, A., et al., Impact of machine learning with multiparametric magnetic resonance imaging of the breast for early prediction of response to neoadjuvant chemotherapy and survival outcomes in breast cancer patients. 2019. 54(2): p. 110–117.

23. Shiralipour, A., et al., Identifying key lysosome-related genes associated with drug-resistant breast cancer using computational and systems biology approach. Iranian Journal of Pharmaceutical Research: IJPR, 2022. 21(1): p. e130342.

24. Khorsand, B., et al., Enhancing ischemic stroke management: leveraging machine learning models for predicting patient recovery after Alteplase treatment. Brain Injury, 2025: p. 1–7.

25. Khorsand, B., et al., Enhancing the accuracy and effectiveness of diagnosis of spontaneous bacterial peritonitis in cirrhotic patients: a machine learning approach utilizing clinical and laboratory data. Advances in Medical Sciences, 2025. 70(1): p. 1–7.

26. Razavi, S.A., et al., Metabolite signature of human malignant thyroid tissue: A systematic review and meta-analysis. Cancer Medicine, 2024. 13(8): p. e7184.

27. Hourfar, H., et al., Machine Learning-Driven Identification of Molecular Subgroups in Medulloblastoma via Gene Expression Profiling. Clinical Oncology, 2025: p. 103789.

28. Sadeghnezhad, E., et al., Cross talk between energy cost and expression of Methyl Jasmonate-regulated genes: from DNA to protein. Journal of Plant Biochemistry and Biotechnology, 2019. 28: p. 230–243.

29. Hesami, Z., et al., Microbiota as a State-of-the-art Approach in Precision Medicine for Pancreatic Cancer Management: A Comprehensive Systematic Review. iScience, 2025: p. 112314.

30. Samandari Bahraseman, M.R., et al., The use of integrated text mining and protein-protein interaction approach to evaluate the effects of combined chemotherapeutic and chemopreventive agents in cancer therapy. Plos one, 2022. 17(11): p. e0276458.

31. Thomassen, I., et al., Peritoneal carcinomatosis of gastric origin: a population-based study on incidence, survival and risk factors. 2014. 134(3): p. 622–628.

32. Kobayashi, D. and Y.J.G.c. Kodera, Intraperitoneal chemotherapy for gastric cancer with peritoneal metastasis. 2017. 20(Suppl 1): p. 111–121.

33. Khorsand, B., et al., Enhancing ischemic stroke management: leveraging machine learning models for predicting patient recovery after Alteplase treatment. 2025: p. 1–7.

34. Payedimarri, A.B., et al., Prediction models for public health containment measures on COVID-19 using artificial intelligence and machine learning: a systematic review. 2021. 18(9): p. 4499.

35. Rescinito, R., et al. Prediction models for intrauterine growth restriction using artificial intelligence and machine learning: a systematic review and meta-analysis. in Healthcare. 2023. MDPI.

36. Zhou, X., et al., Prognostic value of PLR in various cancers: a meta-analysis. 2014. 9(6): p. e101119.

37. Gunaldi, M., et al., Prognostic impact of platelet/lymphocyte and neutrophil/lymphocyte ratios in patients with gastric cancer: a multicenter study. 2015. 8(4): p. 5937.

38. Ishikawa, M., et al., Retrospective comparison of nab-paclitaxel plus ramucirumab and paclitaxel plus ramucirumab as second-line treatment for advanced gastric cancer focusing on peritoneal metastasis. 2020. 38: p. 533–540.

39. Metindir, J., G.J.J.o.c.r. Bilir Dilek, and c. oncology, Preoperative hemoglobin and platelet count and poor prognostic factors in patients with endometrial carcinoma. 2009. 135: p. 125–129.

40. Ejaz, A., et al., Impact of body mass index on perioperative outcomes and survival after resection for gastric cancer. 2015. 195(1): p. 74–82.

41. Chen, H.-N., et al., The impact of body mass index on the surgical outcomes of patients with gastric cancer: a 10-year, single-institution cohort study. 2015. 94(42): p. e1769.

